# Plans to Vaccinate Children Against COVID-19 Among Families Experiencing Homelessness and Other Families in Seattle, WA, October 2020–May 2021

**DOI:** 10.64898/2026.02.05.26344476

**Authors:** Constance E. Ogokeh, Kinda Zureick, Julia H. Rogers, Sarah N. Cox, Amy C. Link, Anne Emanuels, Janet A. Englund, James P. Hughes, Timothy M. Uyeki, Helen Y. Chu, Emily Mosites, Melissa A. Rolfes

## Abstract

**Background:** COVID-19 vaccine hesitancy remains a public health issue despite the fact that vaccines are readily available and recommended for all persons aged ≥5 years in the United States. We aimed to describe parents’ plan to vaccinate their children in two different populations: families in a prospective, longitudinal cohort study and families experiencing homelessness enrolled in cross-sectional surveillance for acute respiratory infections.

**Methods:** Participants were parents/guardians of children aged <18 years, recruited either from a serial cross-sectional homeless study in Seattle-King County, Washington or from a household cohort study in the Seattle Metropolitan area. Participants were surveyed during October 2020—May 2021 about their plans to vaccinate their child against COVID-19. Vaccine plans were described by study population as well as by sociodemographic features and over time.

**Results:** Among parents of 640 children enrolled in the household study surveyed in October 2021, 66% reported planning to vaccinate their child vaccinated against COVID-19 once vaccines became available. This proportion increased slightly over the study period, to 75% in May 2021. In the homeless study, 1284 surveys were collected from parents of 338 children. The proportion of parents of families experiencing homelessness who planned to have their child vaccinated against COVID-19 ranged from 52% in November 2020 to 16% in March 2021.

**Conclusion:** COVID-19 vaccine plan among parents of children experiencing homelessness in Seattle-King County decreased over time, with the majority of parents reporting that they did not plan or were undecided about COVID-19 vaccination for their children by May 2021. Further investigations are needed among families experiencing homelessness to assess vaccine attitudes and perceived barriers to getting their children vaccinated against COVID-19.

**Summary:** Plans to get vaccinated against COVID-19 was less common in children experiencing homelessness and intent decreased over time during the study, whereas vaccination was acceptable in other families in Seattle, WA

## Introduction

While children are less likely than adults to experience severe outcomes of SARS-CoV-2 infection, children contribute to the ongoing transmission and overall burden of coronavirus 19 disease (COVID-19). COVID-19 vaccination remains our best public health tool to mitigate the impact of the pandemic, and vaccinating children may help reduce ongoing transmission.^1–7^ In the United States (U.S.), the Advisory Committee on Immunization Practices (ACIP) currently recommends that all children aged ≥5 years receive a full series and a booster dose of COVID-19 vaccines that have been approved or are under emergency use authorization by the Food and Drug Administration.^8–10^ Several surveys conducted among parents/caregivers in the U.S. have found that roughly half of parents intend to vaccinate their children, but survey results have been heterogeneous with varied attitudes towards COVID-19 vaccines over time.^11–19^ A parent’s intention to vaccinate their children has been associated in several studies with socio-demographic characteristics, such as race and ethnicity, household income, educational attainment, and gender of the parent/caregiver. ^12,20^

A key population missing from many surveys is children experiencing homelessness. On any given night, approximately 170,000 individuals in families experience homelessness in the U.S.; nearly all of these persons reside in sheltered locations and 60% of them are children under 18 years of age.^21^ Children experiencing homelessness may benefit greatly from COVID-19 vaccination as they be more vulnerable to acquiring SARS-CoV-2 infection and developing severe disease.^21–23^ Our primary aim was to measure, among families experiencing homelessness, parents’ intention to vaccinate their children for COVID-19. For comparison, we also measured parental vaccination plans in a longitudinal cohort study of households with school-aged children, being conducted in the same geographic area. Our second objective was to describe changes in plans over time, from the period before vaccines were approved or recommended to when vaccines became available to all adults and were recommended for adolescents, in early summer 2021.

## Methods

### Study design and participants

#### Homeless Shelter Surveillance Study

Beginning in November 2019, all shelter residents (aged ≥3 months) or staff at multiple participating homeless shelters in Seattle-King County were eligible to participate in weekly surveillance for respiratory pathogens; SARS-CoV-2 was included in the protocol starting in April 2020.^24^ Details on participant recruitment have been documented elsewhere.^24^ This analysis focused on children (aged 3 months–17 years) recruited by study staff from three family shelters from November 1, 2020 to May 31, 2021. Interested parents/guardians (hereafter referred as parents) provided consent for their children, and children aged 7–17 years provided assent to participate. A simplified, age-appropriate assent form was provided to children aged 7–12 years, while the parental permission/informed consent form served as the assent form for children aged 13–17. Participation in weekly surveillance included answering a survey questionnaire and providing an anterior nasal swab to be tested for respiratory pathogens. The design of the surveillance was serial cross-sectional, meaning individuals could have enrolled multiple times over the study period (no more than once per week) but no participants were systematically followed longitudinally. Survey data were collected electronically in a secure database via mobile tablet devices at the time of enrollment. Parents of children aged <18 years could choose to complete the survey for their children or allow children aged 12–17 years to complete portions of the survey themselves.

#### Household Study

Households participating in the Seattle Flu Household study were included in this analysis.^25^ Launched in October 2019 with the aim to evaluate the presence and impact of respiratory viruses in families with young children, the Seattle Flu Household study was modified in 2020 to include COVID-19-related questions and testing to detect SARS-CoV-2 infection. During October 2020—March 2021, households were recruited for a new period of study surveillance. Households were recruited from households that had participated in the prior cohort as well as through recruitment flyers distributed throughout the Seattle Public School system. Households were eligible if they were in the Seattle Metropolitan area and had ≥3 members with at least one school-aged child (aged 5–17 years). After obtaining consent (and assent for children aged 7–17 years), participating households were enrolled in active weekly surveillance for acute respiratory illness through June 20, 2021. During the study period, a designated reporter (aged ≥18 years; typically, the parent or guardian, hereafter referred to as parent) in each household completed questionnaires about themselves and their household members.

### Survey questionnaires

Survey questionnaires used in the homeless and household studies included questions on age, sex, race and ethnicity, health insurance status, presence of underlying medical conditions, family income, history of influenza and COVID-19 vaccination, and COVID-19 vaccine plans. Duration of homelessness and history of SARS-CoV-2 testing or exposure were collected only in the homeless study, whereas household structure, adult education, and concern about SARS-CoV-2 infection were collected only in the household study.

In both studies, the questionnaires asked about plans to vaccinate children for COVID-19 using the following question: "*Once a vaccine [When a vaccine] against COVID-19 becomes available to you, do you [or your child] plan to get it?*", with response options of "*Yes*," "*Undecided*," "*No*," and "*Prefer not to say*." The respondents were instructed to answer the questions as they pertained to the child. The option “*Prefer not to say*” was not available in the household study. Those reporting no plans or undecided about getting COVID-19 vaccine were asked to identify the single, primary reason for their lack of plans to be vaccinated from a list of reasons, including: *"Concerns about vaccine safety or efficacy*", "*I need more information about the vaccine*", “*Other reason*”, "*None of the above*", and "*Prefer not to say*" (see Supplementary Table 1).

In the household study, the survey with COVID-19 vaccine plans, as well as receipt of a COVID-19 vaccine, were administered online monthly; the monthly survey conducted in May 2021 was the final survey included in analysis. In the homeless study, the survey with COVID-19 vaccine plans was administered via mobile tablet device whenever an individual participated, which could have been as frequently as weekly.

All characteristics and responses described in this paper, unless otherwise noted, pertain to the child; for example, sex and age represent the sex and age of the child.

### Statistical Analysis

For the homeless study, individuals were able to participate more than once, and participant records were linked over time using available information. Records with the same DOB, sex, and name were assigned the same unique identifier. If a record had the same DOB and sex, but incompatible name spelling, a measure of similarity between the names (Levenshtein distance) was calculated.^26^ The same unique identifier was assigned if the value of similarity was 0.8 or greater (on a scale of 0 to 1). For records with the same name and sex, a one-digit discrepancy in DOB was allowable to assign the same unique identifier. This process was only able to link records within an individual; we were unable to use it to link individuals within family units.

Overall sociodemographic information of participating children was summarized using the most recent survey in the homeless study and the survey conducted at the start of enrollment into the household study. COVID-19 vaccine plans were summarized over time in several ways. First, responses were summarized by month of the study period (for the homeless study, the most recent survey in a month was used when the individual participated more than once). Next, data from each study were divided into two time periods: early (November 2020–January 2021 for the homeless study, October 2020–January 2021 for the household study) and late (February 2021–June 2021). The early period corresponded to the period when COVID-19 vaccines were being developed but not yet recommended for the majority of adults or children and was a period with increased circulation of SARS-CoV-2 in the Seattle area. During the late period (February 2021–May 2021) COVID-19 vaccine was becoming more widely available to the general adult population. For the homeless study, if participants were enrolled more than once during the early time period, the first response about COVID-19 vaccine plans was used in the analysis. For the household study, answers collected at enrollment were used in the early time period. For the late time period, the most recent survey response was used in the analysis. To link surveys from the same children, a unique identifier was used to link records using name, date of birth (DOB) and sex in the homeless study. Finally, individuals who had at least one response during the early and late periods were included in an analysis of change in plans over time, where the first survey response was compared with the last from an individual.

Data analysis and visualizations were conducted using SAS software (Version 9.4) and R Statistical Software (Version 4.0.3). The homeless study did not collect information that permitted direct linkage of survey records within family units; thus, we are unable to account for correlation of responses by family. Due to the difference between the study designs for the homeless shelter and household studies, we did not calculate comparative statistics between the participants enrolled in the two studies.

The homeless shelter and household studies were reviewed and approved by the Human Subjects Division of the University of Washington Institutional Review Board (STUDY00007800 and STUDY00007628, respectively). The CDC relied on the University of Washington reviews. These studies were conducted consistent with applicable federal law and CDC policy (see 45 C.F.R. part 46; 21 C.F.R. part 56).

## Results

### Characteristics of Participating Children

During the study period, 1,284 surveys were collected on 338 children participating in the homeless shelter surveillance study. Children were enrolled a median of two times (range: 1–25) over the study period. A total of 648 surveys were completed during the early time period with half of them completed by December 23, 2020; 636 surveys were completed during the late time period, half of them completed by March 10, 2021. The average age of participating children experiencing homelessness was seven years (range: <1-17 years) and 53% were male. Children were identified as Black or African American (35%), Native Hawaiian or Pacific Islander (17%), multiracial (13%), or White (10%) (Table); 28% were identified as Hispanic. Most enrolled children had experienced homelessness for less than six months (65%), had health insurance (91%) and had been previously tested for SARS-CoV-2 at least once since July 1, 2020 (87%). Among the children previously tested for SARS-CoV-2, 15 (5%) were reported to have tested positive. Half of children (52%) were reported as being vaccinated with or intended to receive the 2020–2021 influenza vaccine.

A total of 345 households (1,363 participants, 640 children) were enrolled in the household study and engaged in weekly symptom surveillance with no loss-to-follow-up. On average, households had 1.9 (range: 1–4) children, and most children enrolled were aged 5–11 years (57%). Children were identified as White (78%) or multiracial (14%), and 93% were non-Hispanic (Table). Almost all the children had health insurance (99%), particularly private health insurance. Most children lived in households where the annual household income was greater than $150,000 (57%) and the highest education level of the adults was an advanced degree (72%). At enrollment, 593 (93%) children were reported to have received or were planning on receiving the 2020–2021 influenza vaccine.

**Table.**
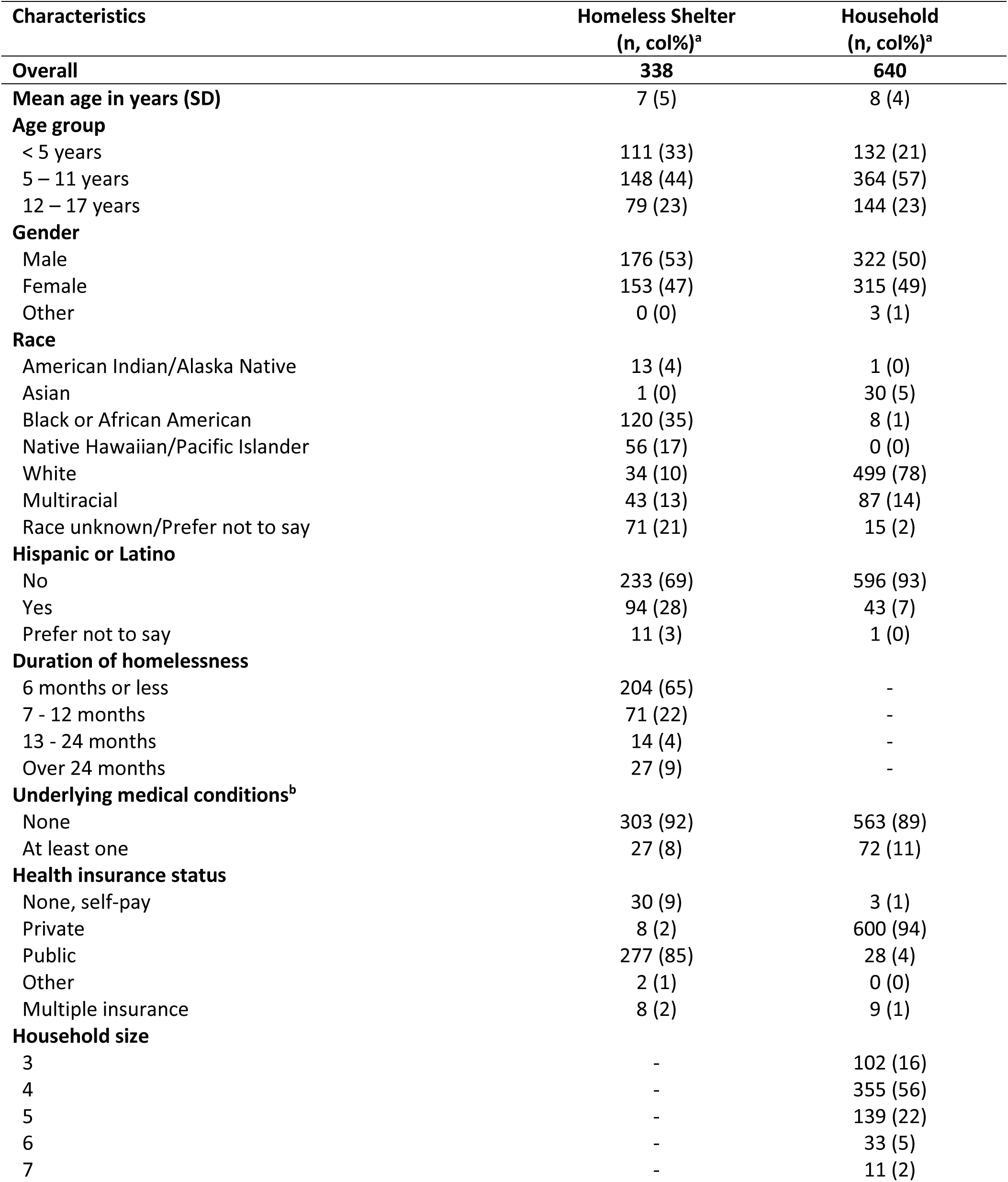
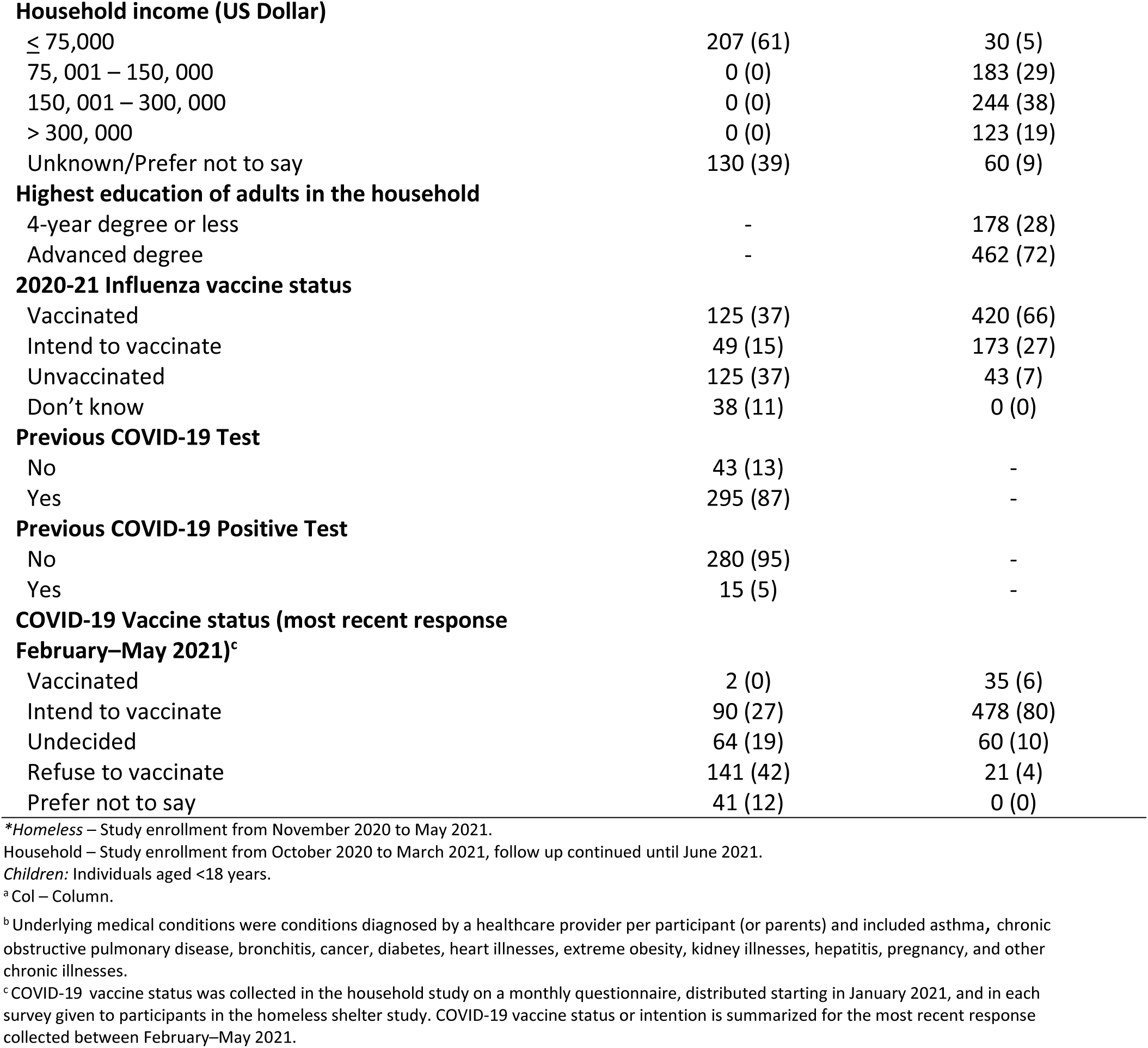
Socio-demographic characteristics, history of COVID-19, history of influenza vaccination, and COVID-19 vaccination plans for children (aged <18 years) experiencing homelessness and children enrolled in a prospective household cohort in Seattle, WA, October 2020–May 2021*

| Characteristics | Homeless Shelter<br>(n, col%)<sup>a</sup> | Household<br>(n, col%)<sup>a</sup> |
| --- | --- | --- |
| **Overall** | **338** | **640** |
| **Mean age in years (SD)** | 7 (5) | 8 (4) |
| **Age group** |  |  |
| < 5 years | 111 (33) | 132 (21) |
| 5 – 11 years | 148 (44) | 364 (57) |
| 12 – 17 years | 79 (23) | 144 (23) |
| **Gender** |  |  |
| Male | 176 (53) | 322 (50) |
| Female | 153 (47) | 315 (49) |
| Other | 0 (0) | 3 (1) |
| **Race** |  |  |
| American Indian/Alaska Native | 13 (4) | 1 (0) |
| Asian | 1 (0) | 30 (5) |
| Black or African American | 120 (35) | 8 (1) |
| Native Hawaiian/Pacific Islander | 56 (17) | 0 (0) |
| White | 34 (10) | 499 (78) |
| Multiracial | 43 (13) | 87 (14) |
| Race unknown/Prefer not to say | 71 (21) | 15 (2) |
| **Hispanic or Latino** |  |  |
| No | 233 (69) | 596 (93) |
| Yes | 94 (28) | 43 (7) |
| Prefer not to say | 11 (3) | 1 (0) |
| **Duration of homelessness** |  |  |
| 6 months or less | 204 (65) | - |
| 7 - 12 months | 71 (22) | - |
| 13 - 24 months | 14 (4) | - |
| Over 24 months | 27 (9) | - |
| **Underlying medical conditions<sup>b</sup>** |  |  |
| None | 303 (92) | 563 (89) |
| At least one | 27 (8) | 72 (11) |
| **Health insurance status** |  |  |
| None, self-pay | 30 (9) | 3 (1) |
| Private | 8 (2) | 600 (94) |
| Public | 277 (85) | 28 (4) |
| Other | 2 (1) | 0 (0) |
| Multiple insurance | 8 (2) | 9 (1) |
| **Household size** |  |  |
| 3 | - | 102 (16) |
| 4 | - | 355 (56) |
| 5 | - | 139 (22) |
| 6 | - | 33 (5) |
| 7 | - | 11 (2) |
| **Household income (US Dollar)** |  |  |
| ≤ 75,000 | 207 (61) | 30 (5) |
| 75, 001 – 150, 000 | 0 (0) | 183 (29) |
| 150, 001 – 300, 000 | 0 (0) | 244 (38) |
| > 300, 000 | 0 (0) | 123 (19) |
| Unknown/Prefer not to say | 130 (39) | 60 (9) |
| **Highest education of adults in the household** |  |  |
| 4-year degree or less | - | 178 (28) |
| Advanced degree | - | 462 (72) |
| **2020-21 Influenza vaccine status** |  |  |
| Vaccinated | 125 (37) | 420 (66) |
| Intend to vaccinate | 49 (15) | 173 (27) |
| Unvaccinated | 125 (37) | 43 (7) |
| Don't know | 38 (11) | 0 (0) |
| **Previous COVID-19 Test** |  |  |
| No | 43 (13) | - |
| Yes | 295 (87) | - |
| **Previous COVID-19 Positive Test** |  |  |
| No | 280 (95) | - |
| Yes | 15 (5) | - |
| **COVID-19 Vaccine status (most recent response February–May 2021)<sup>c</sup>** |  |  |
| Vaccinated | 2 (0) | 35 (6) |
| Intend to vaccinate | 90 (27) | 478 (80) |
| Undecided | 64 (19) | 60 (10) |
| Refuse to vaccinate | 141 (42) | 21 (4) |
| Prefer not to say | 41 (12) | 0 (0) |

<sup>a</sup>Homeless – Study enrollment from November 2020 to May 2021.
Household – Study enrollment from October 2020 to March 2021, follow up continued until June 2021.
Children: Individuals aged <18 years.
<sup>a</sup>Col – Column.
<sup>b</sup> Underlying medical conditions were conditions diagnosed by a healthcare provider per participant (or parents) and included asthma, chronic obstructive pulmonary disease, bronchitis, cancer, diabetes, heart illnesses, extreme obesity, kidney illnesses, hepatitis, pregnancy, and other chronic illnesses.
<sup>c</sup> COVID-19 vaccine status was collected in the household study on a monthly questionnaire, distributed starting in January 2021, and in each survey given to participants in the homeless shelter study. COVID-19 vaccine status or intention is summarized for the most recent response collected between February–May 2021.

### COVID-19 Vaccine Plans

The proportion of respondents planning to vaccinate the child against COVID-19 fluctuated across time in the homeless study, ranging from a high of 52% in November 2020 to a low of 16% in March 2021 (Figure 1). Across the early time period (November 2020–January 2021), 41% of respondents in the homeless study indicated they planned to get their child vaccinated against COVID-19, 36% did not have that plan, and 21% were undecided (Figure 2a). The proportion of parents reporting that they planned to have their child vaccinated was greatest for homeless children who were <12 years old, Hispanic, White, or who had received or planned to receive the 2020-2021 influenza vaccine (Figure 2a). However, during the late time period (February 2021–May 2021), in each socio-demographic category, there was a greater proportion of respondents who did not plan to or were undecided about having the child vaccinated against COVID-19 (Figure 2b). Among the respondents who reported not planning or being undecided during the late period, the primary reason cited for their response was concerns about vaccine safety or effectiveness (21%) and need for more information (18%). Nearly a third (31%) of respondents; however, answered “none of the above” and 28% preferred not to answer the question (Supplementary Table 2).

**Figure 1.**
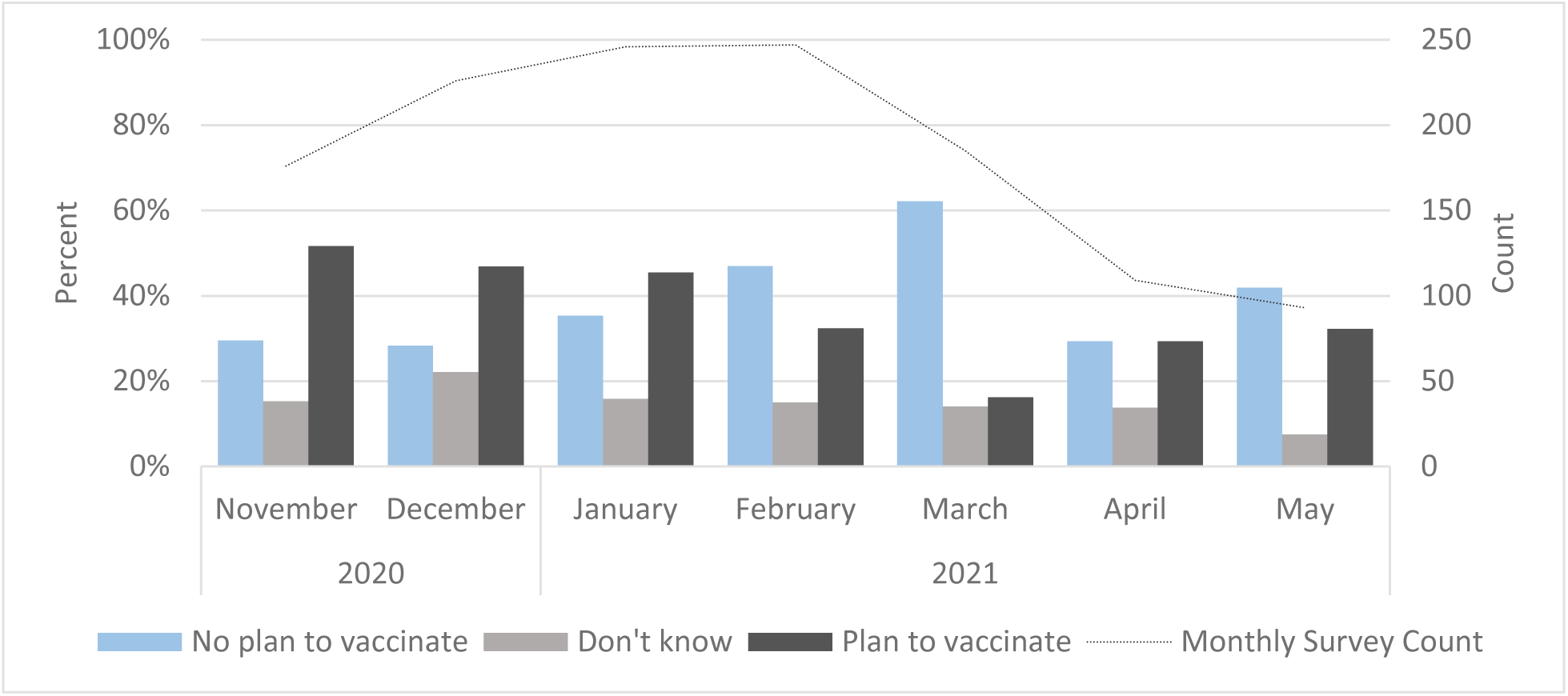
Monthly response to survey asking parents if they plan to vaccinate their child against COVID-19 — families experiencing homelessness, Seattle, WA, November 2020–May 2021

**Figure 2.**
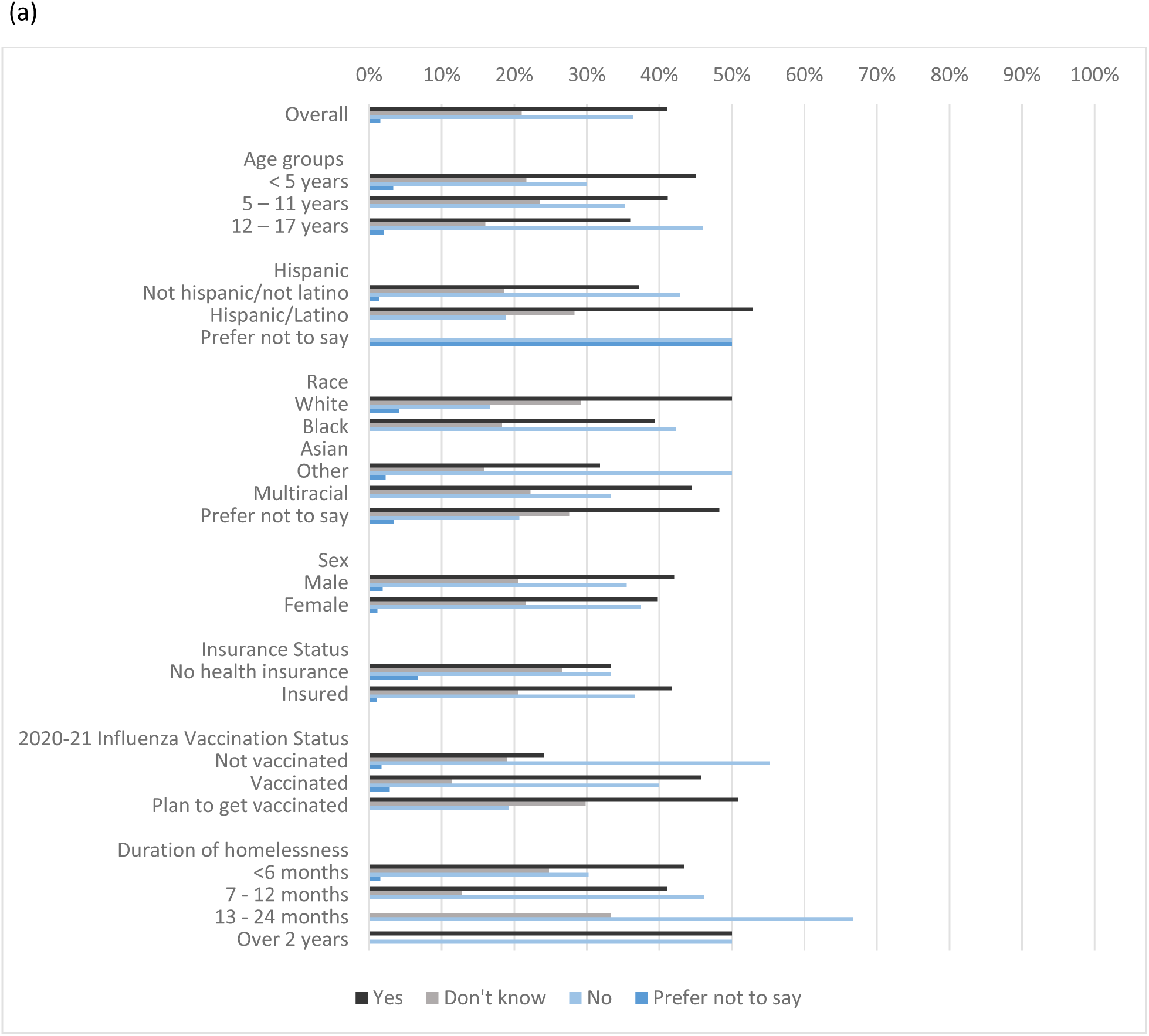
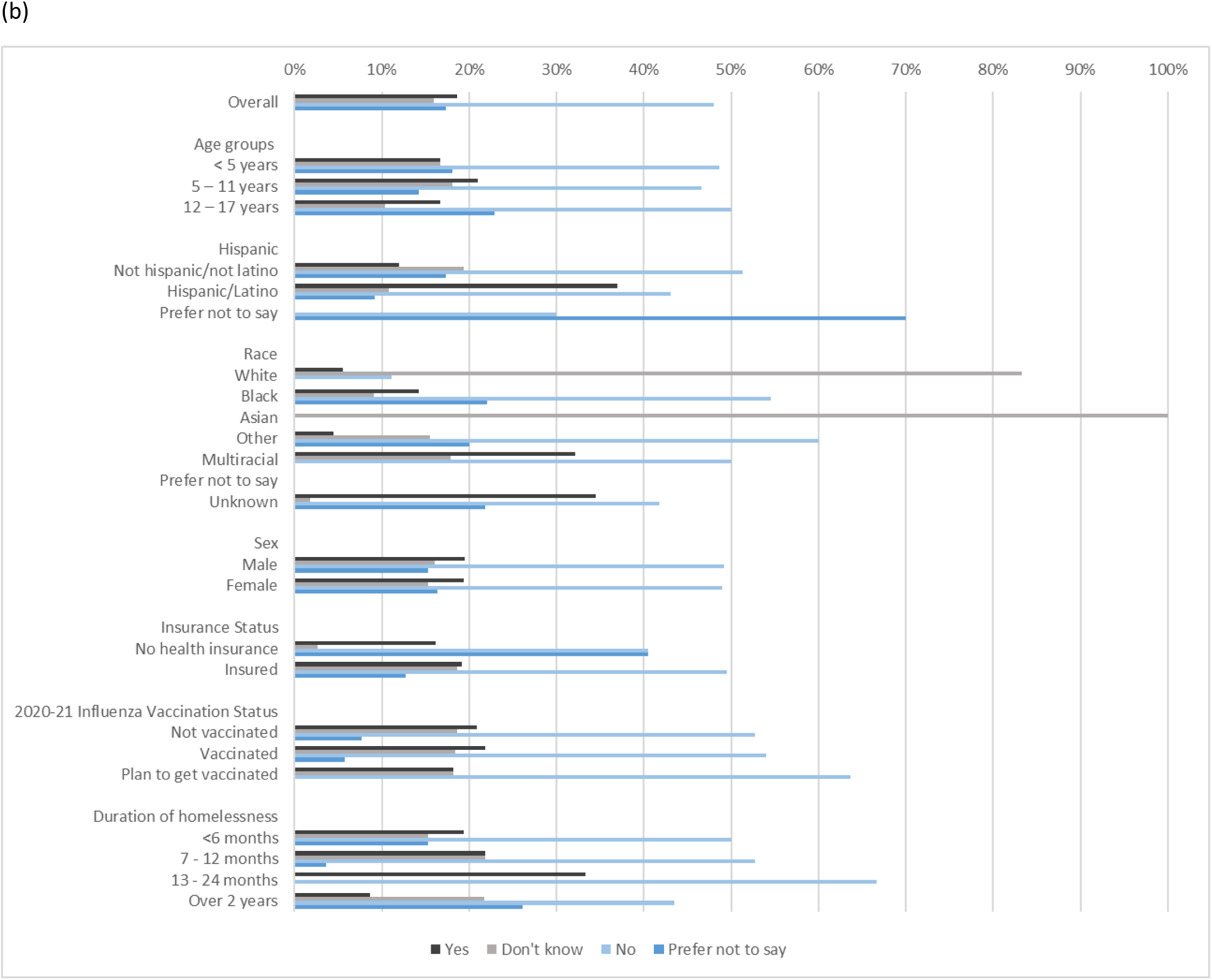
**(a and b).** Responses to survey asking parents if they plan to vaccinate their child against COVID-19, by characteristics of the child — families experiencing homelessness, Seattle, WA. Panel a depicts the earliest responses to the survey during November 2020–January 2021 (N=195) and panel b depicts the most recent survey during February–May 2021 (N=227)

Compared to the homeless study, intention to vaccinate children against COVID-19 in the household study remained consistent across the study period, ranging from 66% planning to vaccinate in October 2020 to a high of 77% planning to vaccinate in May 2021 (Figure 3). In both the early and the late time periods, there were slightly more respondents reporting no plan to vaccinate the child if the child had not received nor intended to receive the 2020-2021 influenza vaccine (Figure 4a and 4b).

**Figure 3.**
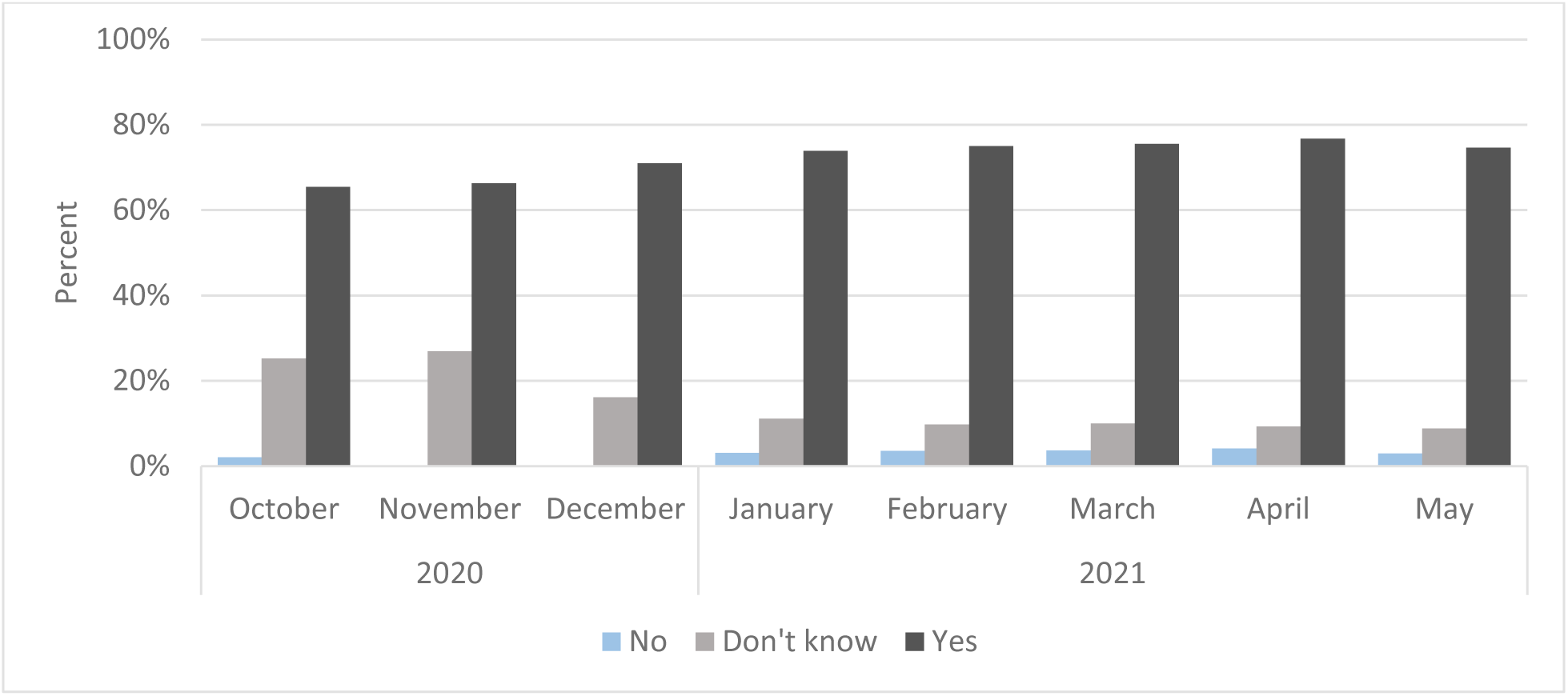
Monthly response to survey asking parents if they plan to vaccinate their child against COVID-19 — prospective household cohort, Seattle, WA, October 2020–May 2021 (N=640)

**Figure 4.**
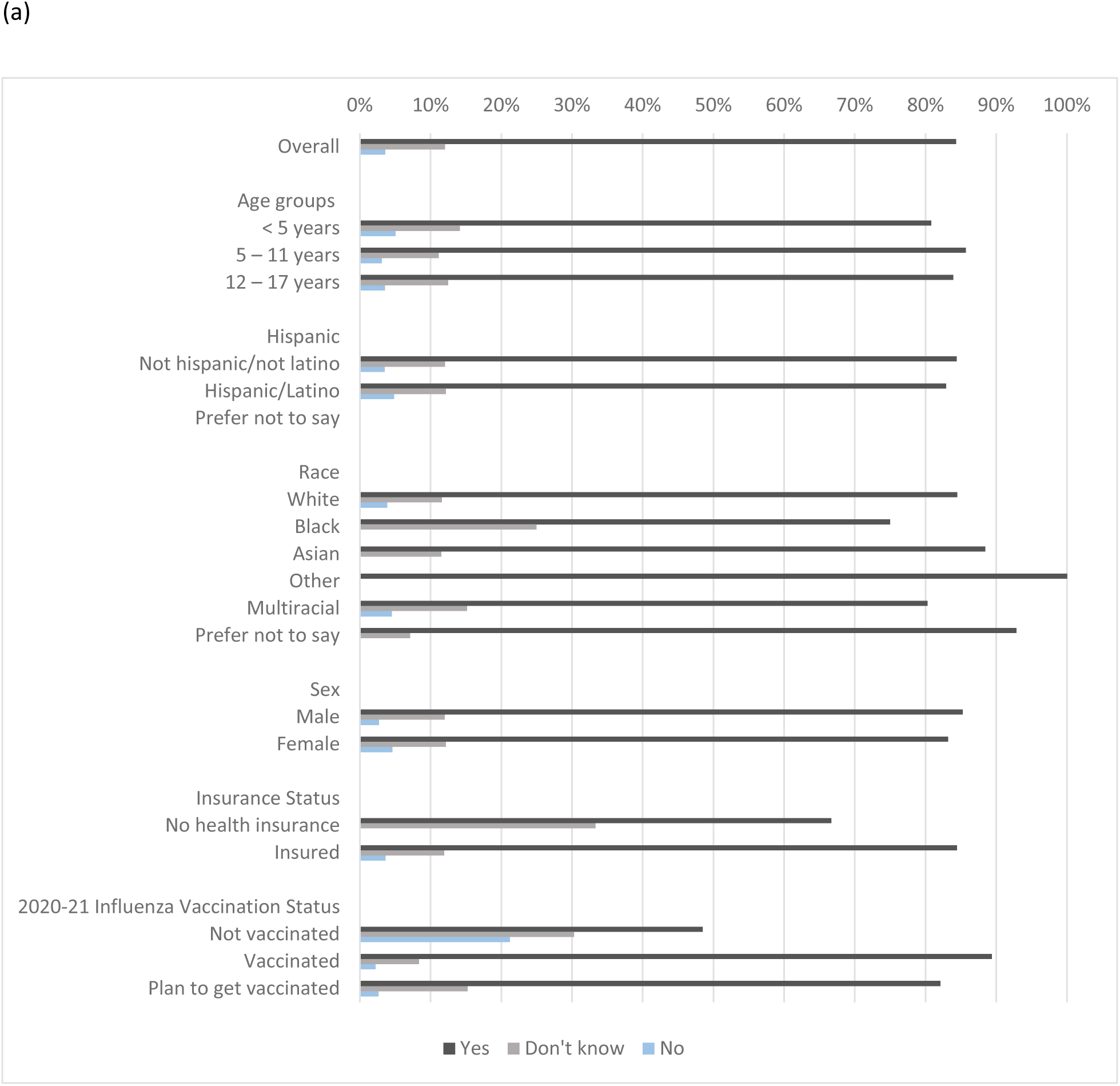
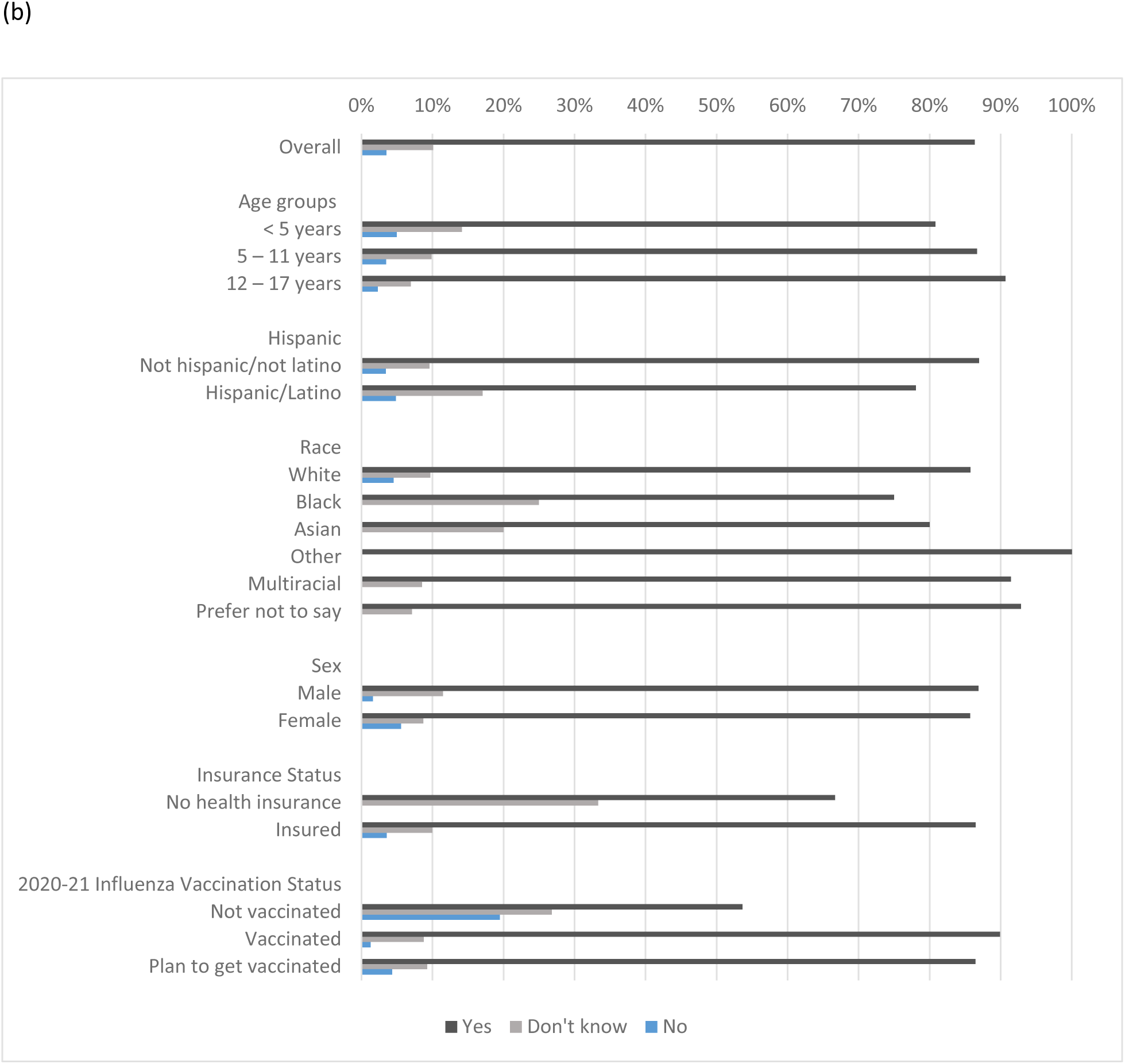
**(a and b)**. Responses to survey asking parents if they plan to vaccinate their child against COVID-19, by characteristics of the child — prospective household cohort, Seattle, WA. Panel a depicts the earliest responses to the survey during October 2020–January 2021 (N=498) and panel b depicts the most recent survey during February–May 2021 (N=594).

### Change in COVID-19 Vaccination Plans Over Time

A total of 84 children were enrolled in the homeless study at least once during both the early and late time periods and had responses to the COVID-19 vaccination questions, allowing comparison of vaccine plans within the same respondents over time. Overall, the proportion of participants with response in both time periods that reported not planning to vaccinate the child increased from 32% in the early time period to 48% in the late time period. Among those who initially did not plan to vaccinate the child, 70% had not changed their response in the most recent survey (Figure 5a). Among those who initially planned to vaccinate the child, 32% maintained that intention, while 45% did not plan to vaccinate their child in their last survey. Among those who were initially undecided about vaccinating the child, parents of 41% children reported they planned to vaccinate their child, while parents of 31% did not plan to vaccinate on their last survey.

**Figure 5.**
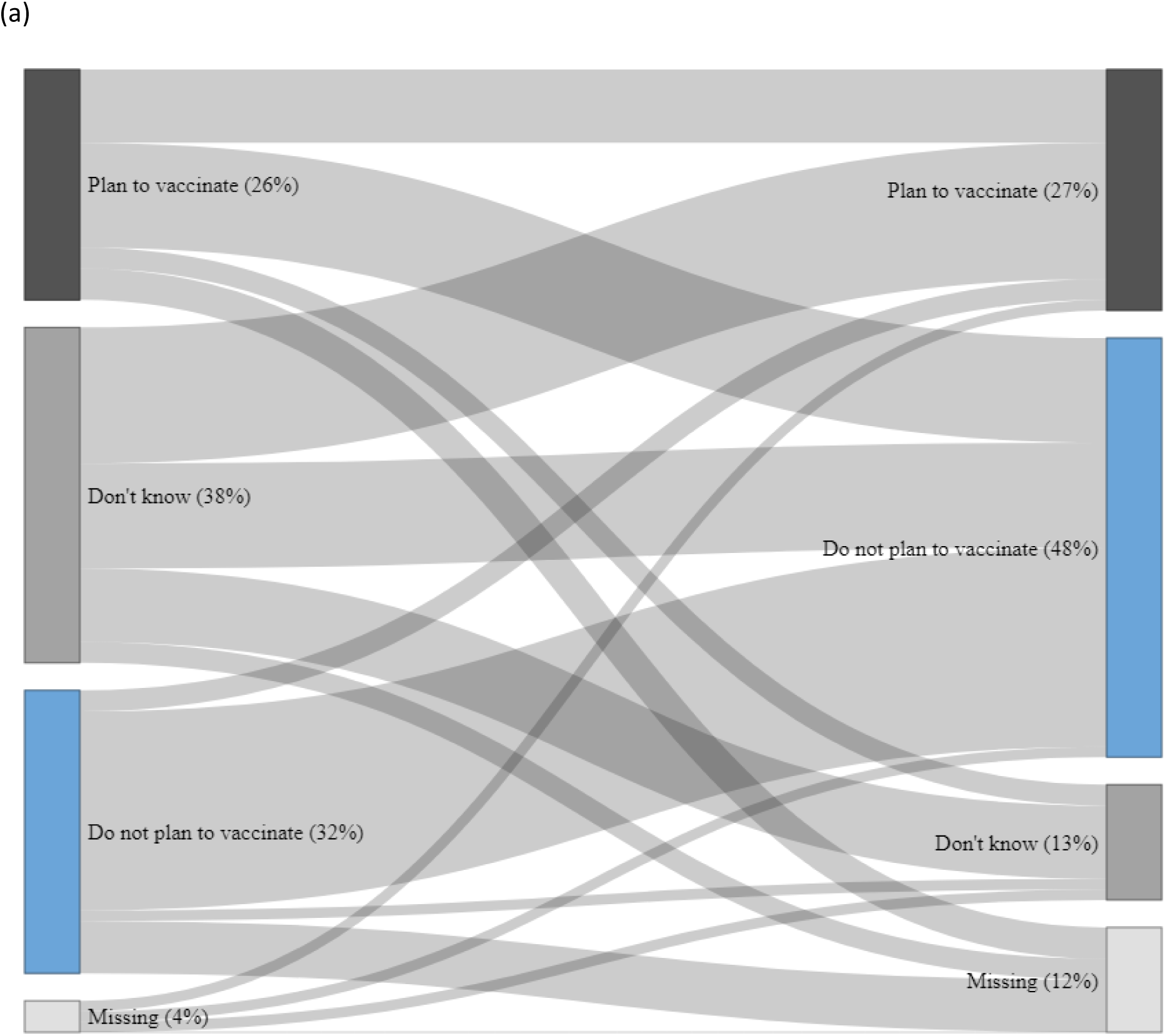
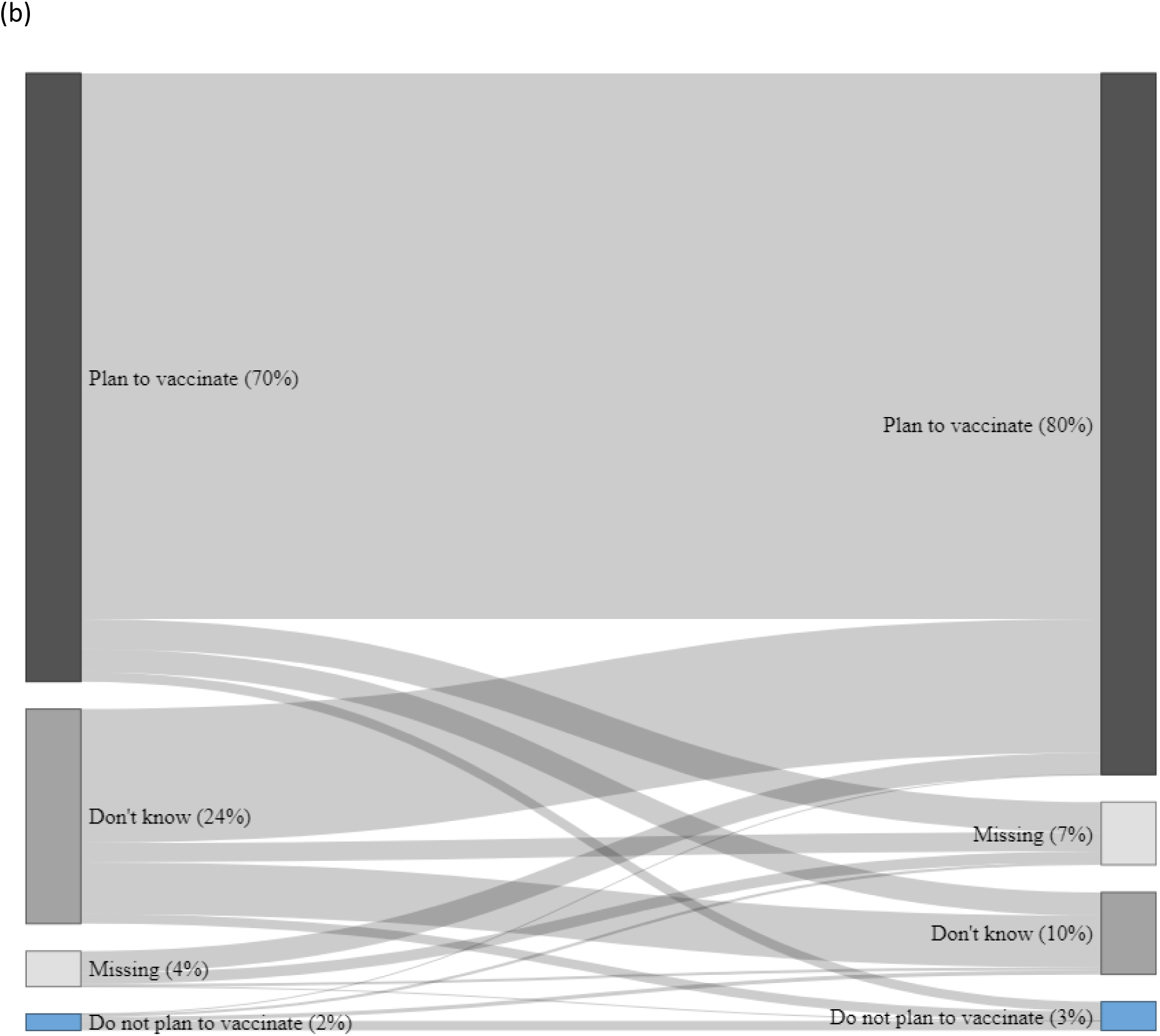
**(a and b).** Changes in parents’ plans to vaccinate their children against COVID-19, Seattle, WA during October 2020–May 2021. Data in panel a come from a serial cross-sectional surveillance study of families experiencing homelessness. Individuals (n = 84) included in panel a participated during both the early and late time periods. Data in panel b come from 640 participants in a prospective household cohort study. In each panel, a participant’s first survey response during October 2020–January 2021 was depicted on the left-hand side of the graph and the most recent survey response during March–May 2021 was depicted on the right-hand side of the graph.

When comparing parental vaccine plans collected during the early to the late time period in the household cohort, parents of 444 (69%) children kept the same intention about their children receiving COVID-19 vaccination, while parents of 132 (21%) changed their intention (Figure 5b). Among the 157 children whose parents indicated at enrollment that they were undecided about their children receiving a COVID-19 vaccination, 98 (62%) indicated in their latest survey that they planned to get them vaccinated, while 38 (24%) remained undecided.

## Discussion

This unique set of parallel studies, conducted in the Seattle metropolitan area during similar time-periods in the COVID-19 pandemic, demonstrated differences in intention toward COVID-19 vaccination of children between two population groups. In households that were generally wealthy, predominately white, and with highly educated parents, planning to vaccinate children was common with little variation between October 2020 and May 2021. However, planning to obtain COVID-19 vaccine was less common among families experiencing homelessness, and we saw an increase in the proportion of parents who did not report planning to vaccinate their children over the study period. The lower number of parents with intention to vaccinate their children while experiencing homelessness should be an area for future investigation, to see if the trends towards vaccine reluctance continued after May 2021, to assess what the drivers of those attitudes may be and understand how those beliefs translated into decisions for or against COVID-19 vaccination in the months after our studies concluded.

Findings from the household cohort in Seattle showed that most parents planned to have their child vaccinated against COVID-19, were similar to data from other published surveys conducted in the U.S.^15,17–19,27^ However, children experiencing homelessness have not been a focus of most COVID-19 vaccination confidence surveys, and there was a surprising shift away from planning to seek vaccination of children. The reasons for the shift in attitudes are likely multifactorial and are worth exploring in future investigations to help shape public health messaging or interventions in this disproportionately affected population. The attitude shift could be related to COVID-19 circulation patterns. The early time period encompassed months during which COVID-19 circulation was increasing in Seattle, including the population of people experiencing homelessness, whereas the later time period had lower virus activity.^28^ Additionally, both studies were conducted prior to widespread local circulation of more transmissible variants of SARS-CoV-2, such as Delta and Omicron.^29^ The shift in attitudes may also have been influenced by news and events surrounding COVID-19 vaccines as the later time period coincided with more reports of adverse events among vaccinated persons, including reports of thrombocytopenia among those who had received adenovirus vector vaccines in the United States and elsewhere, and a brief pause in the administration of the Janssen COVID-19 vaccine.^30–34^

Several socio-economic factors, found in other studies to be correlated with COVID-19 vaccine intentions for children, may also be contributing to the contrast in vaccine intentions that we observed in the two populations in Seattle.^12^ Studies that have assessed predictors of vaccine intentions in the general population have found that the child’s age, parental attitudes towards COVID-19 vaccines, and attitudes towards vaccines for their children in general were predictors of COVID-19 vaccine reluctance in parents or caregivers.^16,35–37^ In general, studies have found that the younger the child and the lower the household income, the more likely a parent will be reluctant to vaccinate their children for COVID-19.^16,35–37^ While the socio-demographic characteristics of the families experiencing homelessness may suggest lower intention during each time period, they do not fully explain the change in attitudes over time.

### Strength and limitations

Our study provides a valuable description of COVID-19 vaccine intentions for children experiencing homelessness over time periods that span vaccine approval and roll-out in the U.S. In addition, we were able to compare vaccine intention among a non-random sample of households during the same time period and in the same geographic area. While the temporal-spatial similarities were a strength, the two populations differed considerably and were not comparable economically or racially. For these reasons, formal statistical comparison would not likely have been informative. Our study has several additional limitations. First, the study designs differed. The household study included systematic longitudinal follow-up of a cohort of households whereas the homeless study had a serial cross-sectional design and repeated observations on the same participant occurred by convenience. These differences could have impacted our conclusions about how COVID-19 vaccine intentions changed over time. Second, there is the potential for selection bias in our analysis of change in intention within individuals over time since repeated participation in the homeless study was not systematic or random. Individuals who participated multiple times in the homeless study may be more health-conscious (because they were seeking testing repeatedly) and were more likely to be homeless for a longer period of time (data not shown). Thus, our conclusions based on this selected group may not be generalizable to the broader population of children experiencing homelessness. Third, the person responding to the survey may be different between the studies. We are more certain that the respondent in the household study was the parent, whereas in the homeless study there may be more responses provided by the child or adolescent. Fourth, while both studies did collect similar socio-demographic information, there were differences, and we were unable to descriptively compare vaccination intention by the educational attainment of the parents/guardians in the homeless study. Finally, the participants in the household cohort may not be reflective of the general Seattle population, making the socio-demographic differences between the household and homeless study participants less easily interpretable. Future investigation should strive to include a broad sample of the population to better assess socio-demographic drivers of vaccine attitudes. Additionally, in-depth qualitative study would help elucidate the beliefs and attitudes towards COVID-19 vaccination in families experiencing homelessness. In the adult homeless population in Seattle, we are conducting qualitative studies; however, these could be expanded to focus on families with the goal to design programs that increase vaccine confidence for children experiencing homelessness.

### Conclusion

We found that a high proportion of homeless parents reported no intent to vaccinate or were undecided about vaccinating their children against COVID-19. Additionally, there was a shift towards greater reluctance over the study period, ending in May 2021. Since completion of the study, children aged 5 years and older are now recommended to receive COVID-19 vaccine and a booster dose. COVID-19 vaccine coverage remains low nationally among children aged 5-11 years.^38,39^ Because vaccine intention is correlated with vaccination, we would expect children experiencing homelessness to have lower uptake. However, the correlation between vaccination intent and actual vaccination is imperfect and future studies should include homeless children or family shelters in periodic COVID-19 vaccine coverage surveys to help guide resource needs.

## Data Availability

All data produced in the present study are available upon reasonable request to the authors.

## Acknowledgements

We would like to thank the children and parents who participated in both the homeless shelter and the household studies. We also acknowledge Seattle Public Schools for allowing us to enroll participants in the household study, as well as the Bush School, and St. Joseph School; and additionally thank the shelter staff, program managers, and study staff at Mary’s Place for their collaboration in recruitment and data collection.

---

**Supplementary Table 1.** COVID-19 related questions collected in the survey questionnaires.

| Questions | Answers | Timing |
| --- | --- | --- |
| What is the primary reason that you are undecided or do not plan to receive a vaccine for the coronavirus? | ▪ Do not have time to get vaccinated<br>▪ I am not worried about getting sick with COVID-19<br>▪ Concerns about vaccine safety or effectiveness<br>▪ Concerns with newness of vaccine or rigor of vaccine testing<br>▪ Want to make sure high-risk individuals get it first<br>▪ I need more information about the vaccine<br>▪ None of the above<br>▪ Prefer not to say | At enrollment |

**Supplementary Table 2.** Additional questions about COVID-19 vaccination asked of participants in the homeless shelter surveillance system —Seattle, WA.

| Question | Answers | Frequency of response<br>(n = 144*) |
| --- | --- | --- |
| What is the primary reason that you are undecided or do not plan to receive a vaccine for the coronavirus? | Concerns about vaccine safety or effectiveness | 30 (21%) |
|  | I need more information about the vaccine | 26 (18%) |
|  | Other reason | 3 (2%) |
|  | None of the above | 45 (31%) |
|  | Prefer not to say | 40 (28%) |

\* 144 respondents reported that they were undecided or did not plan on their child receiving a COVID-19 vaccine when it became available.

